# Assessment of Perioperative Biomedical Equipment Availability, Functionality, and Management Practices Across Rwanda: A Cross-sectional Observational Study

**DOI:** 10.64898/2026.07.07.26357184

**Authors:** Tairu Fofanah, Wubabeba Belay Temesgen, Derbew Fikadu Berhe, Pierrette Ngutete Mukundwa, Anteneh Gadisa Belachew, Naol Belema Gemechu, Gatwiri Murithi, Esperance Mukanahayo, Amanuel Adane Bitew, Amédée Ndizeye, Remy Turc, Senait Bitew Alemu, Jean Baptiste Ntihumbya, Abebe Bekele, Henry E. Rice, Barnabas Alayande

**Author notes:** Corresponding author: Tairu Fofanah, Centre for Equity in Global Surgery, University of Global Health Equity, Kigali, Rwanda. Co-first authors (Tairu Fofanah, Wubabeba Belay Temesgen).

## Abstract

Effective management of biomedical equipment prevents breakdowns, extends equipment lifespan, ensures perioperative safety and cost-efficiency. There are major challenges in managing biomedical equipment, particularly in low- and middle-income countries. This study aimed to assess the availability, functionality, and adherence to maintenance practices of biomedical equipment in operating rooms (ORs) and post-anaesthesia care units (PACUs) across Rwanda. A cross-sectional observational study was conducted at one Level 2 district hospital in each of Rwanda’s five provinces (n=5 sites). Data were collected using three main tools: 1) a medical equipment checklist, 2) a checklist for hospital biomedical management, and 3) direct inspections of selected biomedical equipment. All tools underwent pretesting and face validation with support from biomedical experts prior to data collection in May 2024. Key measures, including the availability and functionality of biomedical equipment, and adherence to maintenance and management practices, were summarised using descriptive statistics. The five hospitals had a total of 16 ORs, 4 PACUs, and 226 pieces of equipment. The overall availability of biomedical equipment was 45%, and the functionality of the available equipment was 96%. The mean adherence rate to national management practices was 66%. The Rwandan government, non-governmental organisations, and hospitals were identified as direct funders of the equipment, accounting for 42%, 12%, and 4%, respectively. However, 42% of the equipment surveyed could not be linked to any of the above sources of acquisition. Among non-functional equipment, 75% was due to a lack of spare parts, while 25% was due to a lack of skills to maintain the equipment. In summary, we found low availability of perioperative biomedical equipment across Rwanda, although the available equipment was highly functional. Adherence to national management practice guidelines was relatively low, threatening the sustainability of functional equipment. We recommend that the government and hospital administrators implement robust, regular auditing systems to ensure proper management of biomedical equipment.

## 1. Introduction

Biomedical equipment (BE) is critical for health system operations, and includes a wide array of devices essential for diagnosis, monitoring, treatment, and rehabilitation. Biomedical equipment requires management throughout its life cycle, such as calibration, repair, user training, decommissioning, and safe disposal, which are typically performed by biomedical engineers and end users [1,2]. The modern healthcare system depends heavily on these BE, making them essential for improving healthcare standards, ensuring prompt patient care, and enhancing patient satisfaction globally [3].

To provide equitable and accessible health services, health facilities must have biomedical equipment that is available in sufficient quantities and of appropriate quality [2]. In low- and middle-income countries (LMICs), much of the BE is donated. Although donated BE can address immediate needs, it often fails to achieve the desired outcomes because it is either not used or underutilised [4]. McDonald *et al*. (2019) found that 40-70% of donated medical devices go unused because they are unsuitable for their intended purposes or because of a lack of staff training and maintenance skills [5]. In West Africa, over 70% of hospital-donated medical equipment did not receive ongoing support for resources and maintenance, often due to ineffective communication between donors and beneficiaries [6].

Health facilities in LMICs often lack basic biomedical equipment [7]. The challenges of managing biomedical equipment in LMICs include equipment unavailability, non-functional equipment, inefficient donation processes, and resource constraints [6,7]. Many countries in sub-Saharan Africa lack robust biomedical equipment management systems, leading to high rates of equipment being out of service [8]. As documented in the Pan-African Surgical Healthcare Forum consensus, African countries recognise the regional challenges in biomedical equipment management and agree on the need to address them collaboratively [9]. Rwanda, like many LMICs, is no exception to these challenges, including difficulty with tracking and maintaining biomedical equipment [10].

Addressing these challenges, the Rwandan Ministry of Health has developed guidelines for medical equipment management practices and recommended equipment package lists for public health facilities [11,12]. However, the extent to which biomedical equipment is available and functional, and to which recommended management practices are used across Rwanda, remains poorly understood.

In this cross-sectional study, we assessed the availability, functionality, and adherence to management practices for biomedical equipment in the operating room (OR) and the post-anaesthesia care unit (PACU) in five level 2 district hospitals across Rwanda. The study used a variety of surveys as well as direct, physical inspection. The findings of this study may provide insights to help policymakers make well-informed decisions about biomedical equipment management practices.

## 2. Methods and Materials

### 2.1. Study design

We conducted a cross-sectional observational study in five level 2 district hospitals across Rwanda to evaluate the availability, functionality, and adherence to biomedical equipment management practices.

### 2.2. Study setting

We purposively selected one district (Level 2) hospital from each of the five provinces in Rwanda (n=5 sites total): Kabgayi Level 2 Teaching Hospital (Southern Province), Kirehe Level 2 Teaching Hospital (Eastern Province), Kibogora Level 2 Teaching Hospital (Western Province), Butaro Level 2 Teaching Hospital (Northern Province), and Kacyiru Level 2 District Hospital (Kigali). We focused on district-level 2 hospitals, as these facilities in Rwanda provide most of the operative care for the country’s population [13].

Our study assessed two areas of biomedical equipment capacity: 1) availability and functionality of biomedical equipment in the OR/PACU, and 2) the implementation of policies and guidelines related to the maintenance and management practices of biomedical equipment in the hospitals.

### 2.3. Data Collection tools

We used three data collection tools: 1) a medical equipment checklist, which primarily addresses equipment availability and functionality, 2) a hospital biomedical equipment management practices checklist, which addresses adherence to implementation of management practices, and 3) direct inspection of selected biomedical equipment, which also addresses functionality.

All survey tools were developed based on Rwanda’s policies and guidelines for biomedical equipment donation and management practices [11] and the Rwanda health service package for public health facilities, which lists 19 equipment types that should be available in every OR and PACU [12] (Supplement Table 1). Biomedical experts guided the development of the tools, face-validated them, and helped in conducting pilot testing to ensure their reliability and validity.

### 2.4. Data collection procedure

Data collection was conducted from 13/05/2024 to 27/05/2024 using the KoboCollect tool by data collectors fluent in both Kinyarwanda and English. Data collectors were assigned to ORs, PACUs, and the biomedical engineering unit across the five hospitals. To ensure data accuracy and completeness, we had biomedical personnel from the study sites present during data collection to provide additional information as needed.

To assess equipment availability, we manually counted and inspected all available equipment in the OR and PACU and confirmed the essential equipment list (Supplement Table 1). We listed any additional equipment not included in the checklist on a separate form. If equipment was physically present and on the essential equipment list, we documented it as available; if it was on the list but not present, we documented it as unavailable.

To assess equipment functionality, we collaborated with biomedical personnel and end users to examine all available equipment. For example, we switched on the suction machine and assessed it to ensure it sucked up fluid in the bowl; if it did, we documented it as functional. Where it was not functional, further questions were asked to determine the reasons for the non-functioning.

To assess adherence to biomedical equipment management practices as prescribed in Rwanda’s guidelines and policies, we reviewed maintenance records against national guidelines. Specifically, we checked whether guidelines were implemented throughout the equipment’s life cycle, including tagging equipment, updating maintenance tags, maintaining maintenance plans and replacement plans, and preparing maintenance reports. 31 criteria were assessed for adherence to maintenance and management practices in accordance with Rwanda’s policies and guidelines (Supplement Table 2). If a criterion was met, we documented it as “YES”; if it was not met, we documented it as “NO”.

### 2.5. Data Analysis

All survey and direct inspection data were cleaned before analysis. We employed descriptive statistics to summarise all key measures. All statistical analyses were conducted using SPSS v. 26 and Excel v. 2021.

Our study had three outcome measures expressed in percentages: (1) Availability of BE, which refers to the total BE physically present in the ORs or PACUs at the time of data collection, against the required equipment as listed in the health service package for public health facilities, (2) Functionality of BE, which refers to the total functional BE in the ORs and PACUs at the time of data collection against the available BE in the same period, and (3) Adherence to BE maintenance and management practices, which refers to the total practice criteria that are 100% met (YES), against the total criteria set:

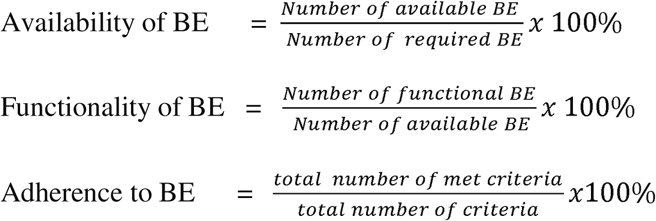

### 2.6. Ethical Declaration

Ethical approval was obtained from the University of Global Health Equity (UGHE) IRB (Ref: UGHE-IRB/2024/309), and a letter of support was received from the Rwanda Ministry of Health; the hospital administration and the biomedical personnel involved in the ORs and PACUs also granted permission. Our research involved facility-based assessment of BE with no protected health information collected. All data were anonymised and stored in compliance with the University of Global Health and the Ministry of Health, Rwanda Institutional Review Board (IRB)-approved protocol, ensuring security and confidentiality throughout the research.

The study did not involve patient interaction. The assessment was purely observational and assessed the availability, functionality and management practices of BE. No patient consent was needed. The study was conducted in accordance with the principles of the Declaration of Helsinki. This study was conducted and summarized in line with the Strengthening the Reporting of Observational Studies in Epidemiology (STROBE) statement (Supplement Table 3).

## 3. Results

We collected data from 16 ORs and 4 PACUs in five district hospitals across Rwanda; these hospitals collectively have 1,327 beds, one biomedical engineer, and eight biomedical technicians (Table 1). Note that two hospitals lacked PACUs at the time of data collection.

**Table 1:**
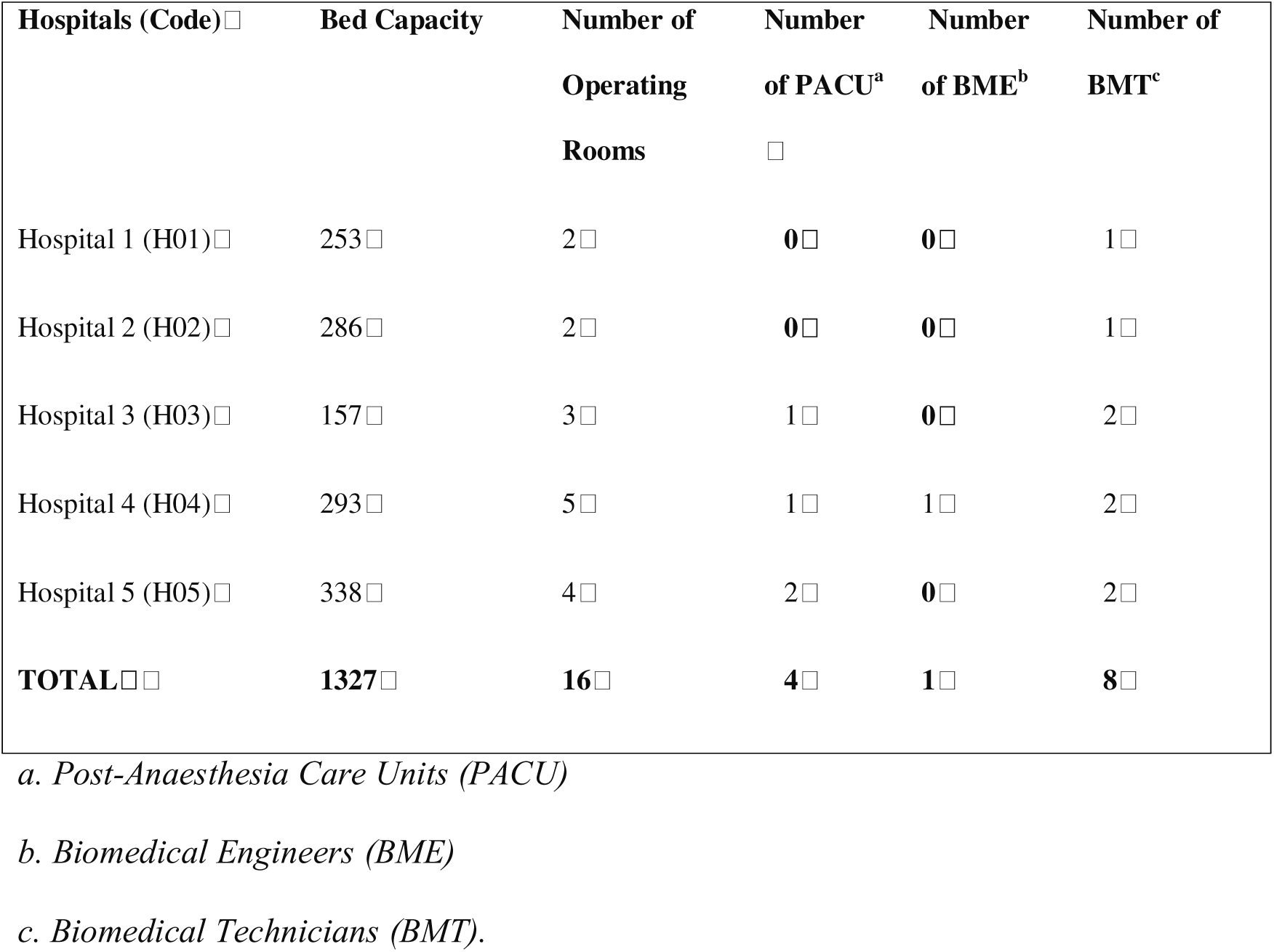
Hospital characteristics in May 2024.

### 3.1. Availability of Biomedical Equipment

In line with the essential medical equipment described in the health service package list for public facilities in Rwanda, the surveyed hospitals had an overall availability of 45% for BE in the OR and PACU. Hospital 1 had the highest availability at 55%, while hospitals 2 and 4 had slightly lower availability, each at 43% (Fig 1).

**Figure 1.** Percentage availability and functionality of biomedical equipment in the Operating Room and Post-Anaesthesia Care Unit across five surveyed hospitals in Rwanda, May 2024

Of the 226 pieces of biomedical equipment available, 42% were from government-supported sources, 12% were donated to hospitals by NGOs, and 4% were purchased directly by hospitals. The remaining 42% of the equipment could not be linked to any of the three sources above.

### 3.2. Functionality of Biomedical Equipment

Of all the available equipment surveyed, 96% were functional. Hospitals 4 and 5 had 100% functionality, while Hospital 2 had the lowest at 83% (Fig. 1). Among the equipment assessed, the most common non-functional types were anaesthesia machines (2 out of 8), infant warmers (1 out of 8), OR lights (3 out of 8), OR tables (1 out of 8) and small oxygen cylinders (1 out of 8). Lack of spare parts accounted for 75% (6 out of 8) of the equipment’s non-functionality, while 25% (2 out of 8) was due to a lack of maintenance technical skills.

### 3.3. Adherence to maintenance and management practices

Of the 31 criteria used to assess adherence to maintenance and management practices for biomedical equipment, hospitals 2 and 4 had the highest adherence at 71% each, hospitals 3 and 5 at 65%, and hospital 1 at 58%. The overall adherence across the hospitals was 66% (Table 2).

**Table 2.**
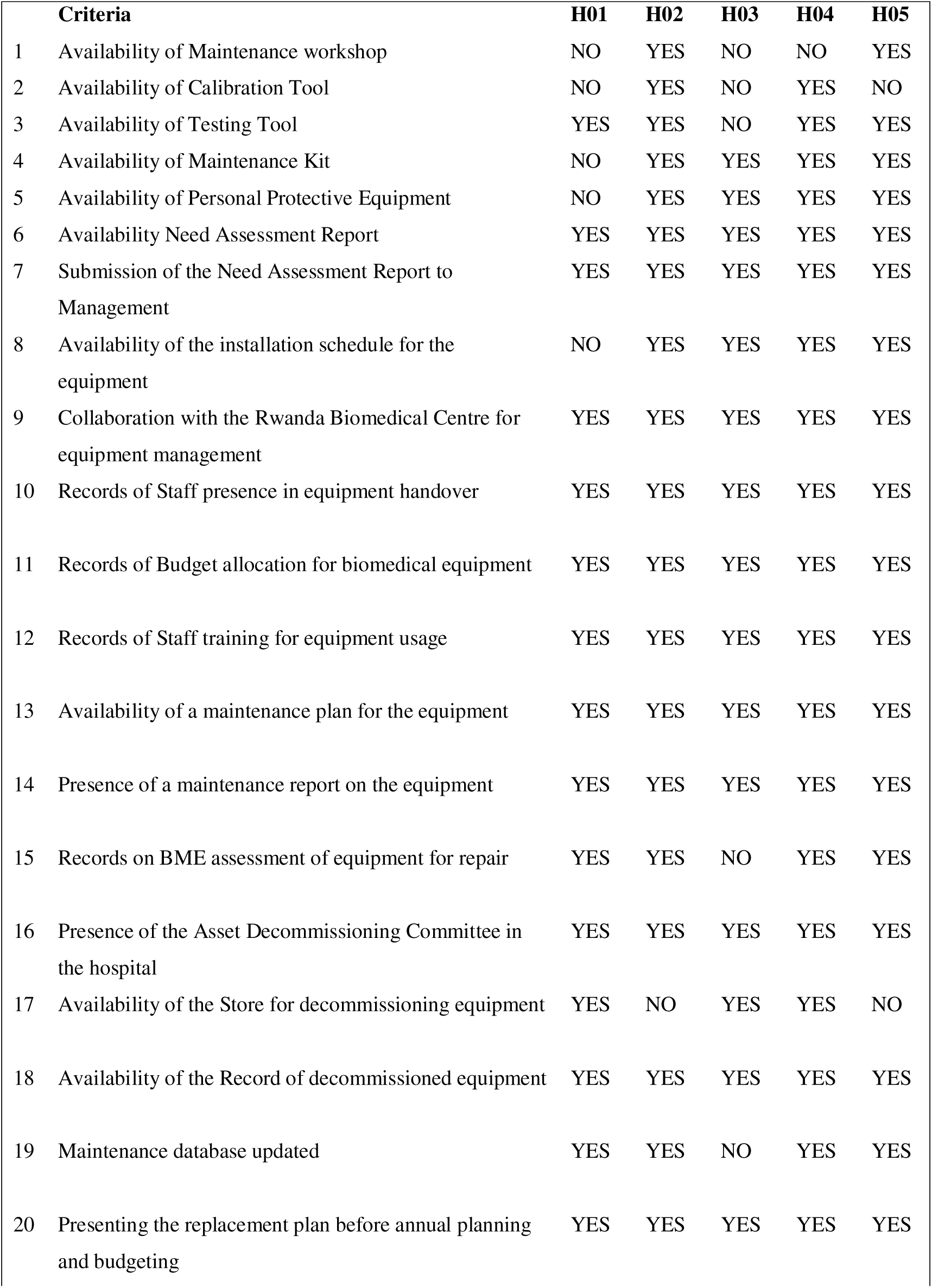
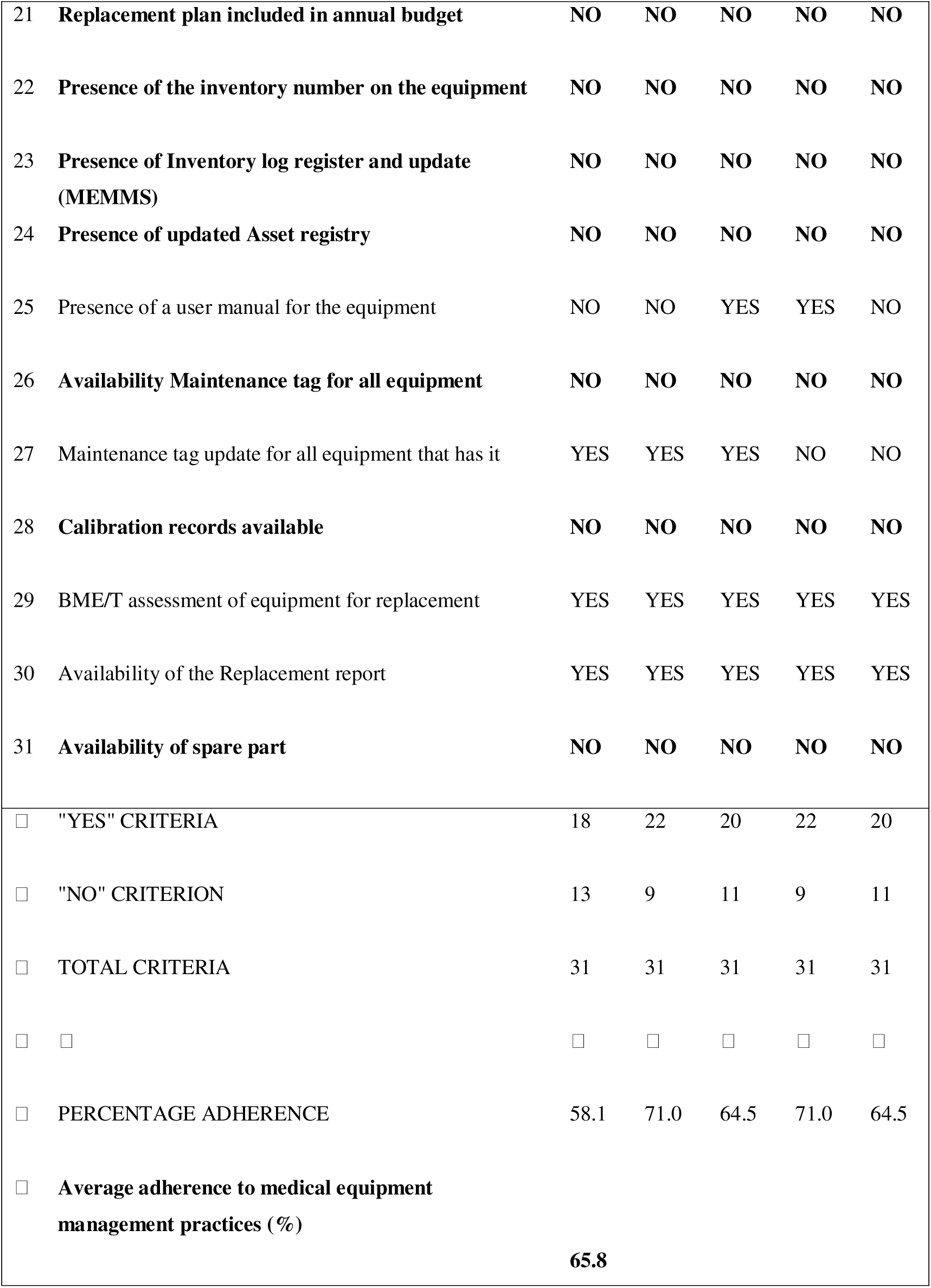
Adherence to criteria for national policies and guidelines on biomedical equipment maintenance and management practices at five district hospitals across Rwanda, May 2024.

|  | **Criteria** | **H01** | **H02** | **H03** | **H04** | **H05** |
| --- | --- | --- | --- | --- | --- | --- |
| 1 | Availability of Maintenance workshop | NO | YES | NO | NO | YES |
| 2 | Availability of Calibration Tool | NO | YES | NO | YES | NO |
| 3 | Availability of Testing Tool | YES | YES | NO | YES | YES |
| 4 | Availability of Maintenance Kit | NO | YES | YES | YES | YES |
| 5 | Availability of Personal Protective Equipment | NO | YES | YES | YES | YES |
| 6 | Availability Need Assessment Report | YES | YES | YES | YES | YES |
| 7 | Submission of the Need Assessment Report to Management | YES | YES | YES | YES | YES |
| 8 | Availability of the installation schedule for the equipment | NO | YES | YES | YES | YES |
| 9 | Collaboration with the Rwanda Biomedical Centre for equipment management | YES | YES | YES | YES | YES |
| 10 | Records of Staff presence in equipment handover | YES | YES | YES | YES | YES |
| 11 | Records of Budget allocation for biomedical equipment | YES | YES | YES | YES | YES |
| 12 | Records of Staff training for equipment usage | YES | YES | YES | YES | YES |
| 13 | Availability of a maintenance plan for the equipment | YES | YES | YES | YES | YES |
| 14 | Presence of a maintenance report on the equipment | YES | YES | YES | YES | YES |
| 15 | Records on BME assessment of equipment for repair | YES | YES | NO | YES | YES |
| 16 | Presence of the Asset Decommissioning Committee in the hospital | YES | YES | YES | YES | YES |
| 17 | Availability of the Store for decommissioning equipment | YES | NO | YES | YES | NO |
| 18 | Availability of the Record of decommissioned equipment | YES | YES | YES | YES | YES |
| 19 | Maintenance database updated | YES | YES | NO | YES | YES |
| 20 | Presenting the replacement plan before annual planning and budgeting | YES | YES | YES | YES | YES |
| 21 | **Replacement plan included in annual budget** | **NO** | **NO** | **NO** | **NO** | **NO** |
| 22 | **Presence of the inventory number on the equipment** | **NO** | **NO** | **NO** | **NO** | **NO** |
| 23 | **Presence of Inventory log register and update (MEMMS)** | **NO** | **NO** | **NO** | **NO** | **NO** |
| 24 | **Presence of updated Asset registry** | **NO** | **NO** | **NO** | **NO** | **NO** |
| 25 | Presence of a user manual for the equipment | NO | NO | YES | YES | NO |
| 26 | **Availability Maintenance tag for all equipment** | **NO** | **NO** | **NO** | **NO** | **NO** |
| 27 | Maintenance tag update for all equipment that has it | YES | YES | YES | NO | NO |
| 28 | **Calibration records available** | **NO** | **NO** | **NO** | **NO** | **NO** |
| 29 | BME/T assessment of equipment for replacement | YES | YES | YES | YES | YES |
| 30 | Availability of the Replacement report | YES | YES | YES | YES | YES |
| 31 | **Availability of spare part** | **NO** | **NO** | **NO** | **NO** | **NO** |
| ☐ | "YES" CRITERIA | 18 | 22 | 20 | 22 | 20 |
| ☐ | "NO" CRITERION | 13 | 9 | 11 | 9 | 11 |
| ☐ | TOTAL CRITERIA | 31 | 31 | 31 | 31 | 31 |
| ☐ | ☐ | ☐ | ☐ | ☐ | ☐ | ☐ |
| ☐ | PERCENTAGE ADHERENCE | 58.1 | 71.0 | 64.5 | 71.0 | 64.5 |
| ☐ | **Average adherence to medical equipment management practices (%)** | **65.8** |  |  |  |  |

All hospitals met 13 management practice criteria in common, which included need assessment, submission of the need assessment to hospital management, collaboration with the Rwanda Biomedical Centre, staff presence during equipment handover, budget allocation for biomedical equipment maintenance, staff training for equipment usage, development of maintenance plan, availability of maintenance reports, presence of asset decommissioning committee, decommissioning records, replacement planning before budgeting, biomedical engineer/technician assessment of equipment, and replacement reports (Table 2).

None of the hospitals met 7 of the 31 practice criteria, including replacement plans in the budget, inventory numbers for all equipment, updated asset registers, maintenance tags on all equipment, calibration records, an inventory log register, updates to the Medical Equipment Management and Maintenance System (MEMMS), and spare-part availability. Only one hospital recorded equipment in the national Medical Equipment Management and Maintenance System (MEMMS), and only two had a designated maintenance workshop.

## 4. Discussion

This study provides a comprehensive assessment of the availability, functionality, and management practices of biomedical equipment in perioperative settings in level 2 district hospitals across Rwanda. We found that less than half of the biomedical equipment recommended by the National Health Service Package guidelines was available, highlighting substantial gaps in perioperative infrastructure. Nevertheless, most of the available equipment was functional, suggesting that maintenance practices are preserving most existing resources. Adherence to national equipment management policies and guidelines was relatively low across facilities. Importantly, our use of direct equipment inspection, in conjunction with structured facility surveys, strengthened the validity of these findings by enabling objective verification of equipment availability, functionality, physical condition, and calibration status. This approach provides a more accurate representation of perioperative equipment capacity than reliance on self-reported data alone.

### 4.1. Availability of BE in the OR and PACU

We found that nearly half of the required biomedical equipment was not present. Given the importance of BE in the delivery of surgical services, the scarcity of essential BE in the OR and PACU may reduce surgical productivity [14]. This gap in available BE can hinder service delivery, increase staff stress when they try to improvise for missing equipment, and increase the risk of medical errors [15]. Our observed availability of BE was lower than that reported by Desalign et al. in a similar study in Ethiopia, which reported 60% availability of required medical equipment in a given facility [2].

Often, even when essential BE was not available, some other expensive surgical equipment was available than required. For example, the National Surgical, Obstetrics, and Anaesthesia Plan of Rwanda (2018-2024) states that there is a need for one backup anaesthesia machine per hospital. However, most surveyed hospitals in the study had one to three backup anaesthesia machines, underscoring the need to improve needs assessment before purchasing equipment to maximise optimal use of resources. A needs assessment is one of the most critical components of medical equipment management, as described in the World Health Organisation document entitled “Needs Assessment for Medical Devices” [16]. A needs assessment can help guide policymakers and other decision-makers in determining appropriate acquisition priorities, storage, and replacement needs.

The government of Rwanda was the largest source of funding for BE, with little funding provided by NGOs. These findings contrast with similar studies in Ethiopia, which found that Tulu Bolo General Hospital received 65% of its equipment as donations [17]. In this study, however, a considerable proportion (42%) of equipment was not linked to any sources of funding. This is comparable to a study in Uganda, where 77% of the equipment was not linked to any source of acquisition [4]. Despite the number of studies reported, most BE in LMICs are donation-based[18,19]. Our study could not align with such reports due to missing information on 42% BE. This discrepancy highlights a major system gap in BE management practices, underscoring the need for proper BE inventory and record management.

### 4.3 Functionality of the BE in the OR and PACU

The majority of the available equipment in the ORs and PACUs was functional. This result is comparable to that of Desalign *et al*., who reported 84% functionality of available medical equipment [2], but differs significantly from Ademe *et al*., who reported that over a third of the medical devices in Jimma zone hospitals in Ethiopia were not functional [20] as well as in Uganda across 9 facilities, which showed 34% equipment was non-functional [4]. A previous study in Rwanda found that high levels of equipment functionality were associated with the presence of trained biomedical technicians in the hospitals [21]. Our study indicates that at least one biomedical technician or engineer is present in every surveyed facility, which could have contributed to the elevated level of equipment functionality.

### 4.4 Adherence to the implementation of maintenance and management practices

We found a relatively low rate (66%) of adherence to national maintenance and management practices in the hospitals surveyed. Adherence to maintenance and management practices, such as end-user training, is crucial at every stage of BE’s life cycle. Findings reveal that all hospitals provide staff training on equipment use, an indispensable component of managing BE [22].

We found that most 75% non-functioning equipment was due to a lack of spare parts. These findings align with those of Tesfaye Geta *et al*. and Desalign *et al*., who reported that a lack of staff training and spare parts impairs equipment functionality [2,23]. Our findings also align with a study in Zambia by Mwanza and Mbohwa, which revealed that spare parts were not documented in the inventory at any of the hospitals and that the facilities purchased spare parts during equipment breakdowns [24]. The lack of spare parts affects the availability and utilisation of equipment, thereby compromising patient care [2].

Ensuring proper maintenance and management practices for BE is crucial for maintaining its functionality and enhancing service delivery. However, hidden costs of maintenance are often overlooked when planning purchases or accepting BE donations. This hidden cost, referred to as the “hippo model” in the healthcare literature, can significantly affect the life cycle of BE if not adequately addressed from the needs assessment stage onward [25].

## 5. Recommendations and regional relevance

Our findings suggest several recommendations for Rwanda and, potentially, for similar health systems across sub-Saharan Africa. First, Ministries of Health and health administrators should prioritise national biomedical equipment lifecycle frameworks that integrate a realistic need assessment, procurement, maintenance, and replacement planning to address persistently low equipment availability. Second, the use of centralised national and regional spare parts supply chains is urgently needed. Third, investment in district-level biomedical engineering capacity must complement end-user training to reduce preventable downtime and strengthen system resilience.

Additionally, countries across sub-Saharan Africa should institutionalise routine audits and digital asset registries to improve accountability, particularly since a substantial proportion of equipment lacks traceable funding sources. Development partners and NGOs should align with national and regional priorities through coordinated donation practices that include maintenance plans, training, spare-part availability, and full-lifecycle support. Strengthening these interconnected system functions across sub-Saharan Africa is critical to sustaining biomedical equipment performance and advancing equitable access to safe perioperative care.

## 6. Limitations of the study

There are several limitations to our analysis. First, as a cross-sectional study that sampled only five hospitals, our study has limited generalizability to other settings across Rwanda or the region. Second, two of the hospitals did not have PACUs at the time of data collection, which also limited generalizability. Third, we focused solely on equipment in ORs and PACUs, limiting our understanding of equipment status in other parts of each hospital. Fourth, equipment functionality was determined by direct inspection and discussion with end users and biomedical personnel, rather than standardised performance testing. Finally, our study could not establish any correlations between functionality and the presence of a biomedical technician due to differences in the number of biomedical personnel across hospitals.

## 7. Conclusions

In summary, we found high functionality of biomedical equipment in the ORs and PACUs in five level 2 district hospitals across Rwanda; however, the overall availability of essential equipment falls well short of the requirements set by the Rwanda health care service package list. Additionally, the inability to track the sources of nearly half of the surveyed equipment poses a significant management challenge.

Moving forward, we recommend that the Rwandan Ministry of Health and other partners focus on improving access to essential, functional BE in ORs and PACUs, including implementing regular audits to ensure hospitals adhere to policies and guidelines for biomedical equipment maintenance and management. Finally, we encourage regular training on the Medical Equipment Maintenance Management System to improve its user-friendliness and adaptability, and we also encourage exploring a mixed-methods approach to understanding the challenges of managing BE.

## Data Availability

The datasets used in this study are available upon reasonable request from the Centre for Equity in Global Surgery at the University of Global Health Equity. The government of Rwanda reserves the right to its data and therefore cannot submit it here. Upon a reasonable request from the centre, it will be available.

## Declarations

### Ethics approval

We obtained ethical approval from the University of Global Health Equity-Institutional Review Board Ref: UGHE-IRB/2024/309 and approval from the Ministry of Health, Rwanda

## Acknowledgements

We want to acknowledge the entire participating hospitals and their staff, including the data collectors, for their participation and Professor Rex Wong of the University of Global Health Equity for his valuable guidance throughout this project and the entire faculty of the Centre for Equity in Global Surgery at the University of Global Health Equity for their invaluable contributions and guidance.

## Authors’ contributions

Tairu Fofanah: Conceptualization, Data curation, Formal analysis, Investigation, Methodology, Project administration, Resources, Validation, Visualization, Writing – original draft, Writing – Review & Editing

Wubabeba Belay Temesgen: Conceptualization, Data curation, Formal analysis, Investigation, Methodology, Project administration, Resources, Validation, Visualization, Writing – Review & Editing

Derbew Fikadu Berhe: Conceptualization, Methodology, Project administration, Resources, Validation, Visualization

Pierrette Ngutete Mukundwa: Conceptualization, Methodology, Project administration, Resources, Validation, Visualization, Writing – Review & Editing

Anteneh Gadisa Belachew: Conceptualization, Methodology, Project administration, Resources, Validation, Visualization, Writing – Review & Editing

Naol Belema Gemechu: Methodology, Resources, Validation, Visualization, Writing – Review & Editing

Gatwiri Murithi: Methodology, Resources, Validation, Visualization, Writing – Review & Editing

Esperance Mukanahayo: Methodology, Resources, Validation, Visualization

Amanuel Adane Bitew: Methodology, Resources, Validation, Visualization, Writing – Review & Editing

Amédée Ndizeye: Methodology, Resources, Validation, Visualization

Remy Turc: Conceptualization, Data curation, Formal analysis, Investigation, Methodology, Project administration, Validation, Writing – Review & Editing

Senait Bitew Alemu: Conceptualization, Data curation, Formal analysis, Investigation, Methodology, Project administration, Validation, Writing – Review & Editing

Jean Baptiste Ntihumbya: Methodology, Supervision, Visualization, Writing – Review & Editing

Abebe Bekele: Conceptualization, Methodology, Supervision, Writing – Review & Editing

Henry E. Rice: Conceptualization, Methodology, Supervision, Writing – original draft, Writing – Review & Editing

Barnabas Alayande: Conceptualization, Methodology, Supervision, Writing – original draft, Writing – Review & Editing

## Supporting Information

**Supplement Table 1**

Essential medical equipment for level two district hospitals OR, described in the health service package list for public facilities in Rwanda.

**Supplement Table 2**

31 Criteria used to assess adherence to biomedical equipment management practices in Rwanda. (crafted from the Rwanda guidelines for medical equipment management policy and guideline).

**Supplement Table 3**

Strengthening the Reporting of Observational Studies in Epidemiology (STROBE) statement.

## References

1. Oshiyama NF, Silveira AC, Bassani RA, Bassani JWM. Medical equipment classification according to corrective maintenance data: a strategy based on the equipment age. Rev Bras Eng Bioméd. 2014; 30:64–9. doi: 10.4322/rbeb.2013.045

2. Desalign D, Keno D, Kaba Z. Assessment of Availability and Utilization of Medical Equipment’s in Nekemte Specialized Hospital, August 2021. Am J Biomed Life Sci. 2023 Feb;11(1):15–21. doi: 10.11648/j.ajbls.20231101.12

3. Arab-Zozani M, Imani A, Doshmangir L, Dalal K, Bahreini R. Assessment of medical equipment maintenance management: proposed checklist using Iranian experience. BioMed Eng OnLine. 2021 May 21;20(1):49. doi:10.1186/s12938-021-00885-5

4. Ssekitoleko RT, Arinda BN, Oshabahebwa S, Namuli LK, Mugaga J, Namayega C, et al. The Status of Medical Devices and their Utilization in 9 Tertiary Hospitals and 5 Research Institutions in Uganda. Global Clinical Engineering Journal. 2022 Mar 1;4(3):5–15. doi:10.31354/globalce.v4i3.127

5. McDonald S, Fabbri A, Parker L, Williams J, Bero L. Medical donations are not always free: an assessment of compliance of medicine and medical device donations with World Health Organization guidelines (2009–2017). Int Health. 2019 Sep 2;11(5):379–402. doi:10.1093/inthealth/ihz004

6. Trunfio T, Baviello D, Perrone A, Formisano R, Donisi L. Medical Technologies Procurement, Management and Maintenance in Developing Countries: The Case of Health Challenges in Africa. In. 2021. p. 793–804. doi:10.1007/978-3-030-64610-3_89

7. Howitt P, Darzi A, Yang GZ, Ashrafian H, Atun R, Barlow J, et al. Technologies for global health. Lancet. 2012 Aug 4;380(9840):507–35. doi:10.1016/S0140-6736(12)61127-1 PubMed PMID: 22857974.

8. Medenou D, Fagbemi LA, Houessouvo RC, Jossou TR, Ahouandjinou MH, Piaggio D, et al. Medical Devices in Sub-Saharan Africa: optimal assistance via a Computerized Maintenance Management System (CMMS) in Benin [Internet]. 2019. doi:10.1007/s12553-018-00283-3

9. Alayande BT, Seyi-Olajide JO, Fenta BA, Ntirenganya F, Obi N, Riviello R, et al. The Pan-African Surgical Healthcare Forum: An African qualitative consensus propagating continental national surgical healthcare policies and plans. Joharifard S, editor. PLOS Glob Public Health. 2024 Nov 12;4(11): e0003635. doi: 10.1371/journal.pgph.0003635

10. Kamanzi JD, Inamarga J, Almasri MW. Medical Equipment Management: A Perispective From Philippines, Rwanda, and Syria. MEDICAL PHYSICS INTERNATIONAL Journal. 2022.

11. MoH Rwanda. Rwanda: Guidelines for Health Care Equipment Donations [Internet]. Ministry of Health, Rwanda; 2017 [cited 2026 Feb 24]. Available from: https://www.medbox.org/document/rwanda-guidelines-for-health-care-equipment-donations

12. MoH Rwanda. Health Service Package for Public Health Facilities [Internet]. 2017. Available from: https://www.moh.gov.rw/fileadmin/user_upload/Moh/Publications/Legal_Framework/Public_health_Facilities_service_packages_in_Rwanda-1.pdf

13. Muhirwa E, Habiyakare C, Hedt-Gauthier BL, Odhiambo J, Maine R, Gupta N, et al. Non-Obstetric Surgical Care at Three Rural District Hospitals in Rwanda: More Human Capacity and Surgical Equipment May Increase Operative Care. World j surg. 2016 Sep;40(9):2109–16. doi:10.1007/s00268-016-3515-0

14. Oosting RM, Wauben LSGL, Groen RS, Dankelman J. Equipment for essential surgical care in 9 countries across Africa: availability, barriers and need for novel design. Health Technol. 2019 May 1;9(3):269–75. doi:10.1007/s12553-018-0275-x

15. Akpor OA, Akingbade TO, Olorunfemi O. Lack of Adequate Equipment for Healthcare – The Agony of Patients and Nurses: A Review. Indian Journal of Continuing Nursing Education. 2023 Jun;24(1):7. doi: 10.4103/ijcn.ijcn_96_21

16. Quintana YL, Cruz LM. Review of management models for needs assessment to acquire biomedical equipment. Health Technol. 2022 Jan 1;12(1):1–8. doi:10.1007/s12553-021-00625-8

17. Kabeta SH, Chala TK, Tafese F. Medical Equipment Management in General Hospitals: Experience of Tulu Bolo General Hospital, South West Shoa Zone, Central Ethiopia. Med Devices (Auckl). 2023; 16:57–70. doi:10.2147/MDER.S398933 PubMed PMID: 36959832; PubMed Central PMCID: PMC10029930.

18. Ojo O, Waheed H. Sustainability challenges in medical equipment donations to low- and middle-income countries. Academia Engineering. 2024 Jun 20;1(2). doi:10.20935/AcadEng6260

19. Marks IH, Thomas H, Bakhet M, Fitzgerald E. Medical equipment donation in low-resource settings: a review of the literature and guidelines for surgery and anaesthesia in low-income and middle-income countries. BMJ Glob Health. 2019;4(5): e001785. doi:10.1136/bmjgh-2019-001785 PubMed PMID: 31637029; PubMed Central PMCID: PMC6768372.

20. Ademe BW, Tebeje B, Molla A. Availability and utilization of medical devices in Jimma zone hospitals, Southwest Ethiopia: a case study. BMC Health Serv Res. 2016 Jul 19;16(1):287. doi:10.1186/s12913-016-1523-2

21. Malkin RA, Whittle C. Biomedical Equipment Technician Capacity Building Using a Unique Evidence-Based Curriculum Improves Healthcare. Journal of Clinical Engineering. 2014 Mar;39(1):37. doi:10.1097/JCE.0000000000000008

22. Doyle PA, Gurses AP, Pronovost PJ. Mastering Medical Devices for Safe Use: Policy, Purchasing, and Training. Am J Med Qual. 2017 Jan 1;32(1):100–2. doi:10.1177/1062860616645857

23. Tesfaye Geta E, Terefa DR, Desisa AE. Efficiency of Medical Equipment Utilization and Its Associated Factors at Public Referral Hospitals in East Wollega Zone, Oromia Regional State, Ethiopia. Med Devices (Auckl). 2023; 16:37–46. doi:10.2147/MDER.S401041 PubMed PMID: 36855514; PubMed Central PMCID: PMC9968427.

24. Mwanza B, Mbohwa C. An Assessment of the Effectiveness of Equipment Maintenance Practices in Public Hospitals. Procedia Manufacturing. 2015 Dec 31; 4:307–14. doi: 10.1016/j.promfg.2015.11.045

25. Corciovă C, Andriţoi D, Fuior R, Luca C. Health Technology Management for Improving the Life Cycle of Medical Equipment. In: 2019 E-Health and Bioengineering Conference (EHB) [Internet]. 2019 [cited 2026 Jun 22]. p. 1–4. Available from: https://ieeexplore.ieee.org/abstract/document/8969908 doi:10.1109/EHB47216.2019.8969908

